# Linking dynamic connectivity states to cognitive decline and anatomical changes in Alzheimer’s disease

**DOI:** 10.1101/2024.10.25.24316107

**Authors:** Jacopo Tessadori, Ilaria Boscolo Galazzo, Silvia F. Storti, Lorenzo Pini, Lorenza Brusini, Federica Cruciani, Diego Sona, Gloria Menegaz, Vittorio Murino

## Abstract

Alterations in brain connectivity provide early indications of neurodegenerative diseases like Alzheimer’s disease (AD). Here, we present a novel framework that integrates a Hidden Markov Model (HMM) within the architecture of a convolutional neural network (CNN) to analyze dynamic functional connectivity (dFC) in resting-state functional magnetic resonance imaging (rs-fMRI). Our unsupervised approach captures recurring connectivity states in a large cohort of subjects spanning the Alzheimer’s disease continuum, including healthy controls, individuals with mild cognitive impairment (MCI), and patients with clinically diagnosed AD. The framework successfully identified distinct brain states associated with different clinical stages of AD, demonstrating a progressive reduction in functional flexibility as disease severity increased. Specifically, we observed that patients with AD spend more time in brain states dominated by unimodal sensory networks, while healthy controls exhibited more transitions to polymodal, cognitively demanding states. Importantly, the fraction of time spent in each state correlated with cognitive performance and anatomical atrophy in key regions, providing new insights into the disease’s progression.

Our findings suggest that the disruption of dynamic connectivity patterns in AD follows a two-stage model: early compensatory hyperconnectivity is followed by a decline in connectivity organization. This framework offers a powerful tool for early diagnosis and monitoring of AD progression and may have broader applications in studying other neurodegenerative conditions.

## Introduction

Alzheimer’s disease (AD) is the most common cause of dementia, characterized by a progressive decline in cognitive function. Early disruptions in neural connectivity, particularly in functional brain networks, have been identified as key indicators of the disease’s onset and progression. Restingstate functional magnetic resonance imaging (rs-fMRI) has emerged as a valuable tool for studying these connectivity alterations ***van den Heuvel and Sporns (2019***), allowing researchers to investigate how different brain regions communicate even in the absence of external stimuli. The analysis of functional connectivity (FC) from rs-fMRI can provide insights into how neurodegenerative processes unfold, offering potential biomarkers for early diagnosis and tracking disease progression. Traditional methods of analyzing FC, such as seed-based correlation and independent component analysis, typically assume that connectivity is static throughout the fMRI scan. However, recent advances in neuroscience emphasize the dynamic nature of brain connectivity, where distinct patterns of FC emerge and dissipate over time ***Favaretto et al. (2022); Fiorenzato et al. (2019); de Vos et al. (2018***). Dynamic functional connectivity (dFC) captures this temporal variability, offering a more nuanced view of brain network alterations that may be critical in understanding diseases like AD. Previous research has shown that dynamic changes in connectivity states may reflect compensatory mechanisms in early stages of AD, followed by a breakdown of connectivity organization as the disease progresses.

Despite these advancements, current methods for analyzing dFC often rely on disjointed processing pipelines, where FC computation, dimensionality reduction, and state classification occur in separate stages. This fragmentation can hinder the optimization of dFC models, limiting their ability to provide a comprehensive understanding of connectivity patterns. To address this, we introduce a novel unsupervised deep learning framework that integrates a Hidden Markov Model (HMM) within a convolutional neural network (CNN). This end-to-end architecture simultaneously optimizes both functional connectivity state identification and sequence likelihood, providing a robust, dynamic representation of brain activity.

The use of Hidden Markov Models (HMMs) for the analysis of FC states is not novel, in itself, but past approaches typically relied on disjointed processing pipelines. In contrast, our method incorporates HMMs directly within a deep neural network, ensuring optimal clustering of brain states in a single architecture. For example, HMMs have been used to characterize brain state dynamics in PTSD subjects ***Ou et al. (2015***), to estimate both static and abruptly changing patterns in children and adults ***Zhang et al. (2019***) and, more recently, to infer emotions evoked with naturalistic stimuli ***Tan et al. (2022***). The closest approach to the one we describe here is perhaps that by Suk et al. ***Suk et al. (2016***), where a deep neural network is employed for dimensionality reduction, followed by a HMM for temporal cluster identification. Nevertheless, in all of the models described above, HMMs are applied after a rather lengthy and separated data preparation phase, whose disjointed application could potentially hinder a comprehensive optimization of the process. In our knowledge, the present work is the first attempt to provide the integration of a HMM directly in a deep neural network.

We apply this framework to a large cohort of subjects spanning the Alzheimer’s disease continuum, from healthy controls to individuals with mild cognitive impairment (MCI) and clinically diagnosed AD. By identifying recurring connectivity states, our model reveals significant alterations in brain network flexibility associated with AD progression. Specifically, we observe that patients with AD exhibit prolonged engagement in brain states dominated by unimodal cortices, while healthy individuals show more frequent transitions to polymodal, cognitively demanding states. These findings align with the growing body of research suggesting reduced metabolic activity and impaired network integration in AD.

Furthermore, our results demonstrate that the fraction of time spent in specific connectivity states correlates with clinical measures of cognitive function and anatomical atrophy in key brain regions. This dynamic, state-based approach to functional connectivity offers a new perspective on the early detection and monitoring of AD, with potential implications for other neurodegenerative conditions. Our method not only identifies clinically relevant connectivity patterns but also provides a scalable tool for exploring brain network dynamics in a wide range of neurological disorders.

**Figure 1.**
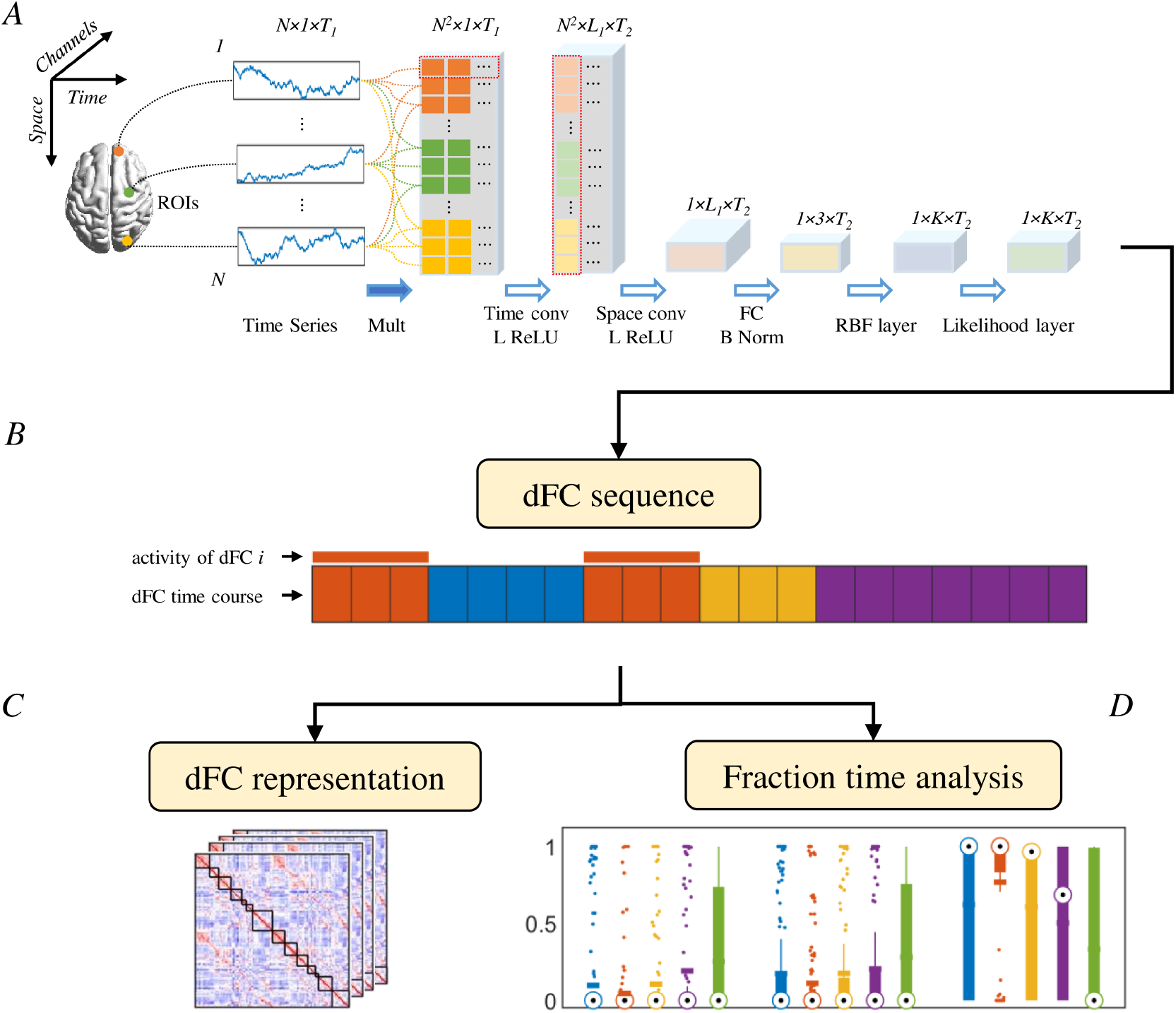
A. Architecture of the proposed framework for dFC construction and analysis using fMRI data. The network receives as inputs the pairwise products of fMRI time series from all ROIs, then two convolutional layers (time conv and space conv) compute time-weighted correlations and provide a low dimensional representation of input data. The last two layers (RBF and expectation layer) reconstruct the most likely sequence of states in the input time series. B. From the output of the neural network, a discrete time series is derived, in which each value indicates the most likely active state for that time point. C. An approximate representation of the different states is provided. D. The fraction time of the identified recurring states is calculated and investigated as a possible marker of AD progression, also in association with cognitive/clinical scores and neurodegenerative trajectories.

## Results

### Experimental settings

We performed two experiments, on data from 1) HC and full-blown AD subjects and 2) in the whole AD clinical spectrum (SMC, eMCI, lMCI, and AD). With the first experiment, we aimed to identify a subset of dFCs (dynamic functional connectivity states) common in both HC and AD subjects. In the second set of experiments, we intended to demonstrate how the distribution of fraction times for the identified dFCs changed according to the clinical condition (from HC to AD) and whether it related to behavior and neurodegenerative trajectories. All experiments were unsupervised and training was performed on all relevant data at once (i.e., no cross-validation was performed).

**Figure 2.**
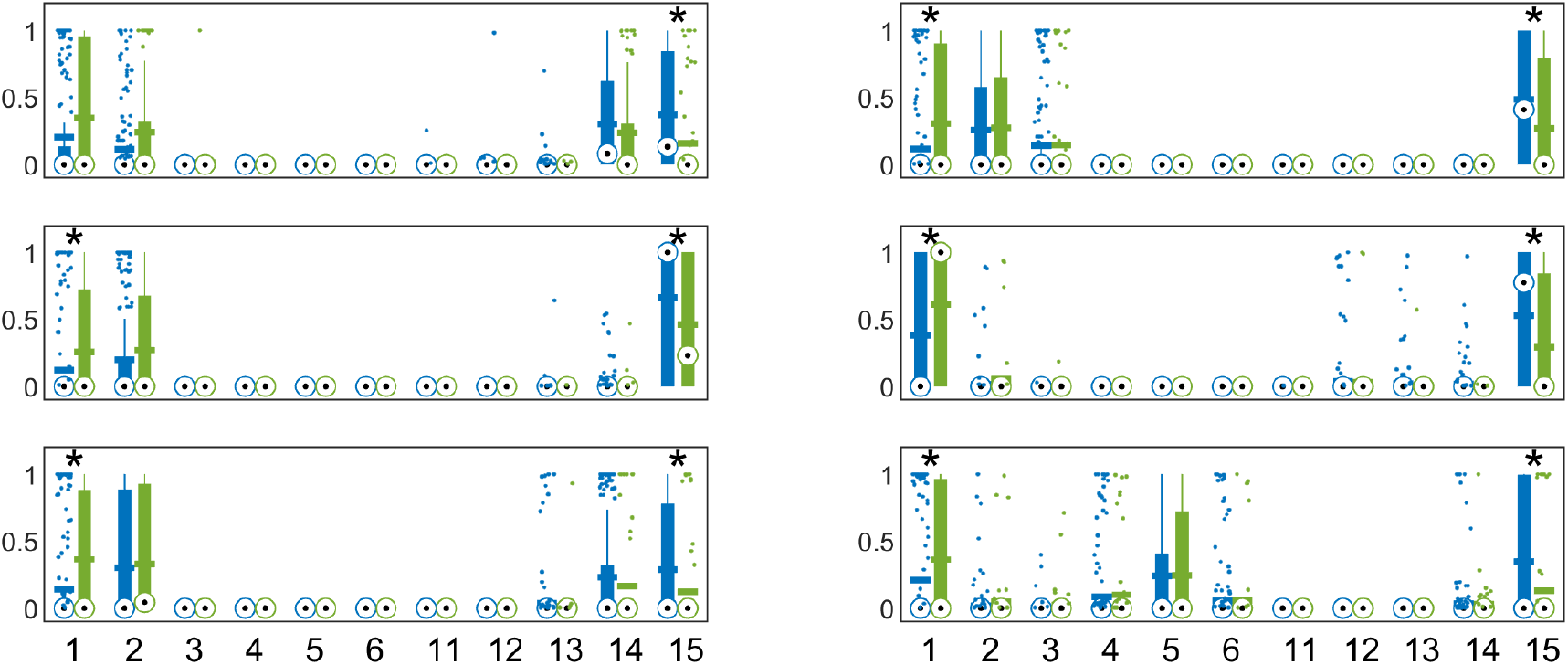
Fraction times for all 6 training runs, with distributions relative to HCs in blue and ADs in green. The symbol * indicates p-value <0.05 for Wilcoxon’s test for equal median, with Benjamini-Hochberg correction for *n*=66 multiple comparisons, 226 HCs, 67 ADs. Circled black dots indicate median values of each distributions, horizontal bars indicate the mean, while bottom and top edges of the box extend to the 25^th^ and 75th percentiles, respectively. Whiskers extend to the most extreme data points not considered outliers (plotted individually as small dots). The representation order is based on the difference between the mean fraction times of HCs and ADs, with ADs over-represented on the left and HCs over-represented on the right. Only 11 distribution pairs are represented, as, after sorting, 4 dFCs positions are never occupied.

### Population comparisons: HC and AD

To demonstrate the variability and reproducibility of the results obtained from the adopted model, we conducted six separate training runs on the HC-AD subset using identical inputs: the only difference among them is, therefore, the random initialization of the model. Each subplot in fig. 2 shows the fraction time of the dFCs identified for a sample training run of the HC-AD experiment. The model is designed to identify a maximum of 15 distinct dFCs. It is evident how, with the adopted settings, no more than 8 dFCs are ever observed in a single training run. Furthermore, some of the most commonly occurring states appear with significantly different fraction times between HC and AD subjects, as confirmed by statistical analysis (Wilcoxon’s tests for equal medians with Benjamini-Hochberg correction, corrected *p*-values smaller than 0.05 are marked with asterisks).

Comparing the results of the six runs can be challenging due to the unsupervised nature of the experiments, which led to the arbitrary numbering of identified dFCs. To facilitate comparison, dFCs have been sorted according to the gap in mean fraction time between HC and AD populations. Using this sorting approach, only 11 out of the possible 15 ranks have at least one non-empty instance across the six training runs. The remaining four positions have been omitted from fig. 2.

With the precautions outlined above, it was possible to verify whether, in different training repetitions, the attributions to a given dFCs were consistent with one another: we performed a permutation test. For dFCs 1, 2 and 15, the counts of matching time volumes in all the six training runs (e.g. for subject 1, time point 1 is always identified as dFCs 1) are at least one order of magnitude higher than the average across permutations (table 1), suggesting that the observed match counts are significantly higher than what can be expected by random chance (*p*-value ≈ 0 for dFCs 1, 2 and 15, Wilcoxon signed rank for median of permutations equal to observed value). Therefore, we can state that these three dFCs were reliably identified in all training runs, thus we decided to focus on them in the rest of the paper. It is worth pointing out that the existence of a significant number of time volumes systematically attributed to the same dFCs across training runs implied that the ranking of dFCs (at least for states 1, 2 and 15) was always consistent.

**Table 1.** Counts of time volumes across all subjects attributed to same dFCs in all training runs in observed data and permutation tests

| dFCs | Observed | Mean perm. | SD perm |
| --- | --- | --- | --- |
| 1 | 4586 | 8.6 | 3.0 |
| 2 | 130 | 1.0 | 1.0 |
| 3 | 0 | 0 | 0 |
| 4 | 0 | 0 | 0 |
| 5 | 0 | 0 | 0 |
| 6 | 0 | 0 | 0 |
| 11 | 0 | 0 | 0 |
| 12 | 0 | 0 | 0 |
| 13 | 0 | 0 | 0 |
| 14 | 0 | 0 | 0.2 |
| 15 | 5638 | 208.1 | 14.3 |

#### Dynamic functional states and their representation

In order to provide a representation of the dFCs, we identified the time volumes that were systemically attributed to dFCs 1, 2 and 15 in all experimental repetitions and constructed “traditional” dynamic FC matrices from the relevant sections of the individual recordings. More in detail, we set the size of each rectangular window to 60 s, so every single FC matrix is constructed with the same data that the convolutional section of the model uses, even though with different time weights. Representations of each dFCs were then obtained by taking the median value from the resulting FCs across all HC/AD subjects. The results are displayed in fig. 3, where the FC matrices are reported along with the information of Yeo’s 7 networks ***Schaefer et al. (2018***) (visual - VIS, somatomotor - SMN, dorsal-attention - DAN, ventral-attention - VAN, limbic - LIM, frontoparietal - FPN and default mode network - DMN, for left and right hemispheres).

### Population comparisons: All groups

In our second experiment, the model was trained on subjects from the 5 available populations. Given the stability of the results observed in the previous experiment, we deemed it sufficient to perform only one repetition of model training. We sorted the order of the identified dFCs exactly as in the previous case. Fraction times are reported in fig. 4, panel A. It is possible to appreciate a state-clinical gradient in terms of fraction times. Specifically, AD, and lMCI patients spent more time in state 1 and state 2, showing a decrease for state 15. HC subjects show a divergent pattern, with the highest fraction of time in state 15 and a steady decrease in states 1 and 2. Notably, the SMC groups showed a similar trend compared to HC, with even an higher amount of time spent in state 15 compared to the healthy counterpart. Table 2 displays the *p*-values for Kruskal-Wallis tests for equal distributions performed on the state-specific fraction times, with Dunn-Sidak correction for multiple comparisons.

**Table 2.** *p*-values of fraction times comparisons in different populations, Kruskal-Wallis test for equal distributions, Dunn-Sidak correction for multiple comparisons (*n*=10).

| Group 1 | Group 2 | dFC 1 | dFC 2 | dFC 15 |
| --- | --- | --- | --- | --- |
| HC | SMC | 0.86 | 0.51 | **0.03** |
| HC | eMCI | 1.00 | 0.83 | 0.98 |
| HC | IMCI | **<1e-2** | 1.00 | 0.13 |
| HC | AD | **<1e-2** | 0.56 | **<1e-4** |
| SMC | eMCI | 0.53 | **0.03** | **<1e-2** |
| SMC | IMCI | **<1e-2** | 0.40 | **<1e-4** |
| SMC | AD | **<1e-3** | **0.03** | **<1e-4** |
| eMCI | IMCI | 0.14 | 0.99 | 0.84 |
| eMCI | AD | **0.03** | 1.00 | **<1e-2** |
| IMCI | AD | 0.99 | 0.87 | 0.11 |

#### Clinico-anatomical interplay with dFCs

In order to investigate whether the identified dFCs have a behavioral meaning, we tested for a possible association between state-specific fraction times and three different clinical scores widely used in AD: Alzheimer’s Disease Assessment Scale (ADAS11), Mini-Mental State Examination (MMSE) and Clinical Dementia Rating Scale Sum of Boxes (CDRSB). The rationale behind this analysis is that if a specific brain state is neurobiologically relevant, its occurrence (as measured by fraction time) might be associated with cognitive performance and clinical outcomes.

HC and AD subjects were involved in the sorting of states, thus in their numbering. In order to eliminate any possible confounding effects on the results of the association analysis, this has been restricted to MCI and SNC populations. For each dFCs, MCI and SMC subjects have been divided in two sub-populations: one for subjects with fraction times for a given dFCs greater or equal to 0.5 and the second for subjects showing that dFCs for less than half of the recording time. As the different sub-populations had different age ranges, the different groups obtained in the previous step have been age-corrected as detailed in the Materials and Methods section in order to minimize any possible impact of age on the outcome. The resulting distributions of the clinical scores in the two sub-populations are represented in fig. 4, panel B. In order to test for a possible association, a Wilcoxon test was conducted for each pair, with a Benjamini-Hochberg correction for multiple comparisons (*n*=9). Resulting *p*-values are presented in table 3. The statistical analyses highlighted a significant association between the fraction times of dFCs 1 and 15, and all the three clinical scores. Subjects with higher fraction time for dFCs 15 showed the best clinical scores compared to the cohort expressing less rs-fMRI frames for this dFCs. This result was reversed for state 1, while state 2 proved significantly associated only with the CDRSB score. Similarly, we explored a possible association between fraction times and anatomical variations in MCI and SMC groups. In this case, we tested whether the sub-populations identified by fraction time thresholding were characterized by significant differences in the relative volumes of hippocampus, amygdala and cerebellum cortex. The former two regions are known to be vulnerable to AD pathology, whereas the latter is generally preserved even in the latest stages (***Braak and Braak (1991); Thompson et al. (2003); Frisoni et al. (2002***)). Fig. 4, panel C displays average relative volumes for the obtained sub-populations, after age-correction. Table 4 reports the *p*-values of a Wilcoxon test for different median conducted on each pair of subpopulations, with a Benjamini-Hochberg correction for multiple comparisons. Fraction times of all the considered states displayed an association with relative volumes of hippocampus and amygdala, while differences for the cerebellum cortex resulted not significant at the 0.05 level.

**Figure 3.**
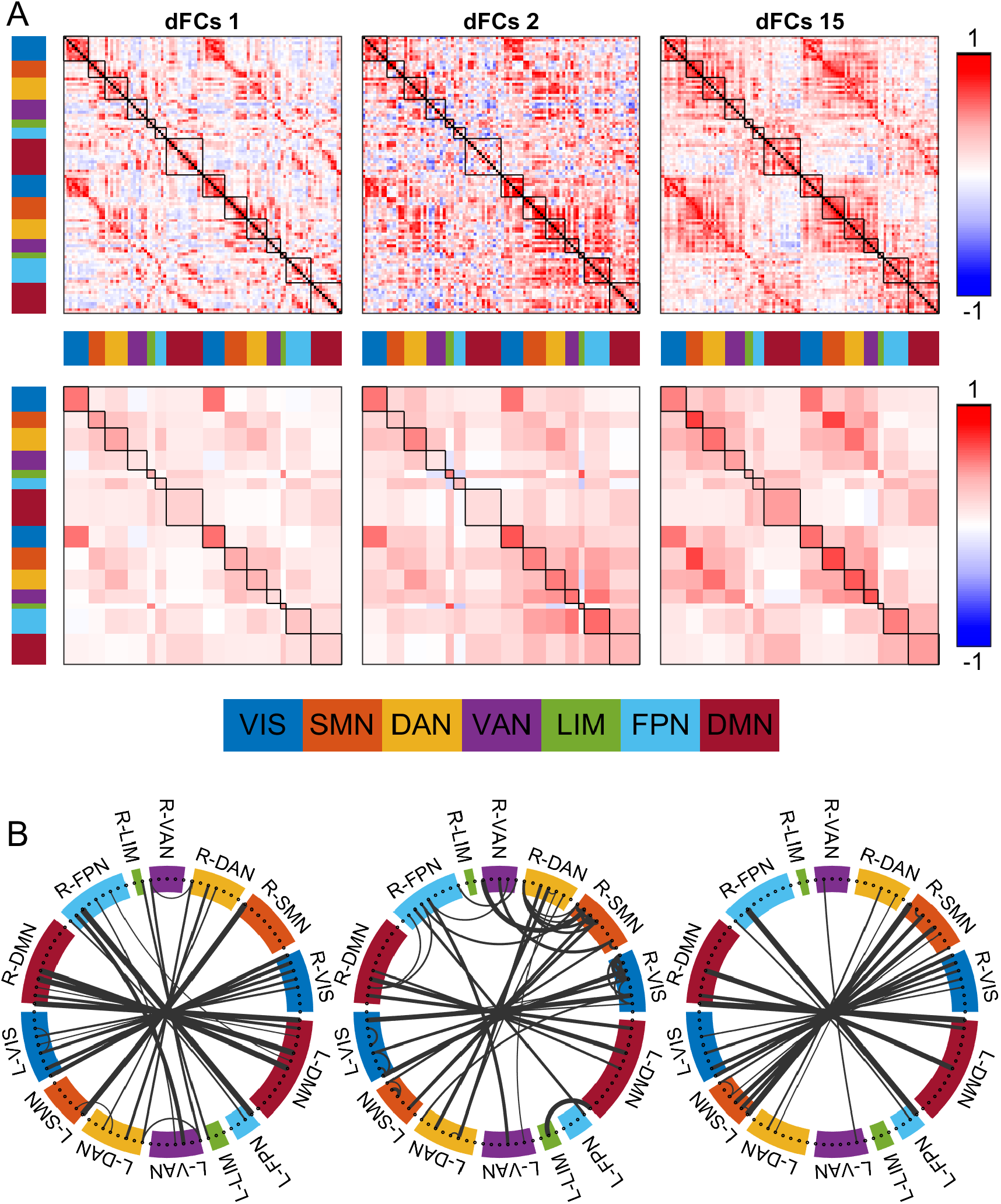
Panel A. Median of FC matrices attributed to dFCs 1, 2, and 15 (respectively for left, middle and right columns) over all training repetitions. The color bar on the right is the same for all panels, while the multicolor bars on the left and bottom of each image provide labelling for the different networks covered by the Schaefer atlas with 7 networks. Bottom row panels display the same data but averaged over intra/inter-networks (excluding values on the main diagonal). Panel B. Connectograms of median FC matrices over all training repetitions. Only connections with values above 0.85 are represented, with line thickness proportional to the connection strength.

**Table 3.** *p*-values of population comparisons of clinical scores in sub-populations divided by state fraction times (Wilcoxon’s test with Benjamini-Hochberg’s correction for n = 9 comparisons).

| Score | dFCs 1 | dFCs 2 | dFCs 15 |
| --- | --- | --- | --- |
| ADAS11 | **<1e-3** | 0.08 | **<1e-4** |
| MMSE | **<1e-4** | 0.11 | **<1e-4** |
| CDRSB | **<1e-3** | **0.05** | **<1e-4** |

**Table 4.** *p*-values of population comparisons of relative region volumes in sub-populations divided by state fraction times (Wilcoxon’s test, Benjamini-Hochberg’s correction for n = 9 comparisons).

| Score | dFCs 1 | dFCs 2 | dFCs 15 |
| --- | --- | --- | --- |
| Hippocampus | **<1e-2** | **0.01** | **<1e-3** |
| Amygdala | **0.01** | **0.01** | **<1e-3** |
| CerebellumCortex | 0.41 | 0.47 | 0.25 |

**Figure 4.**
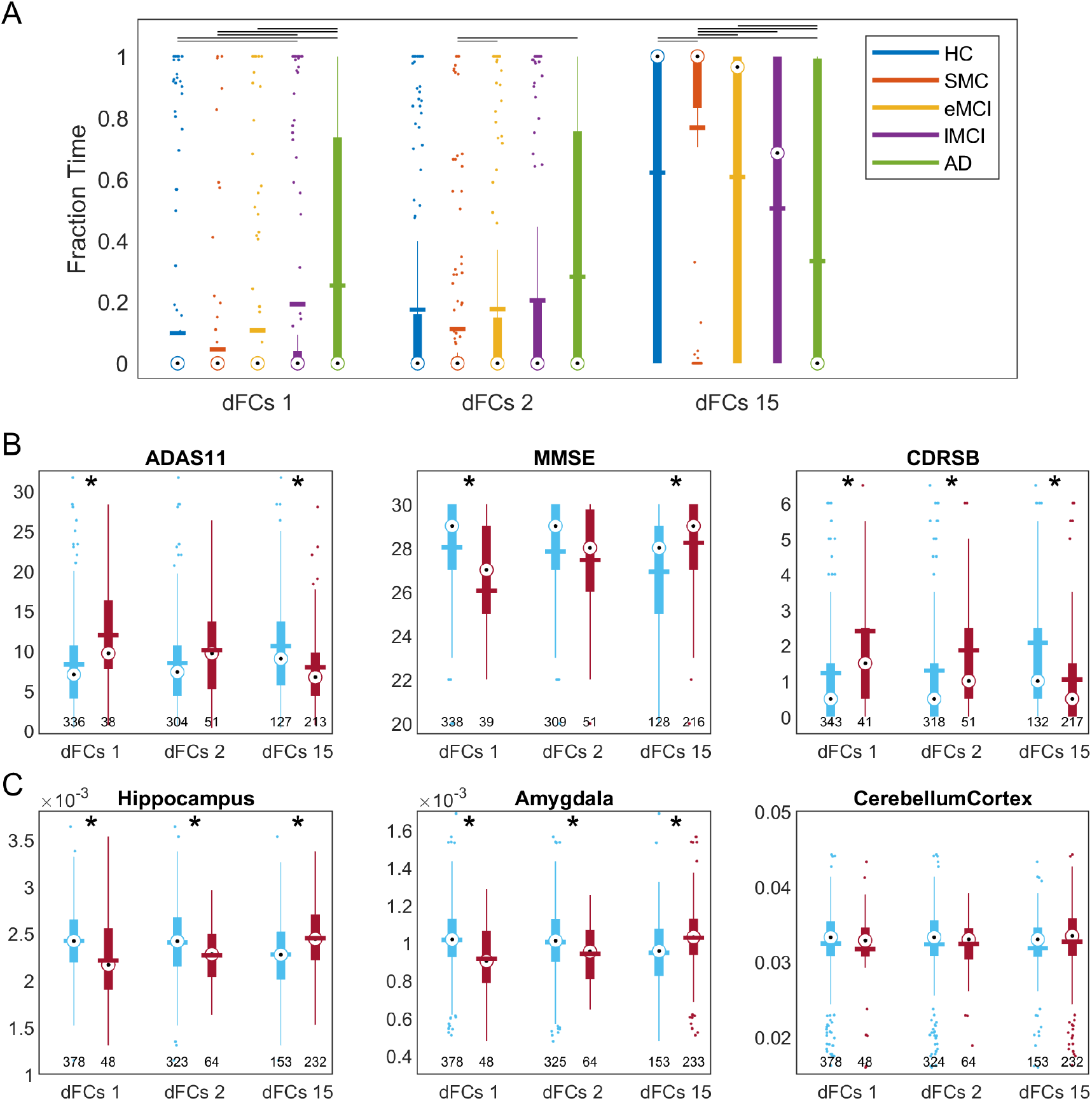
Panel A. Fraction times of the three recurring dFCs, with distributions separated by subject group. The significance between each pair of groups has been tested with Kruskal-Wallis test for equal distributions, with Dunn-Sidak correction to account for the multiple comparisons. The horizontal black lines in the top part of the figure link distributions for which *p*-values are smaller than 0.05. *n*=226 HCs, 160 SMCs, 144 eMCIs, 130 lMCIs, 67 ADs. Boxplots are built as in fig. 2. Panel B. Distribution of three clinical scores in the two sub-populations identified according to the fraction times of each dFCs, with blue and red bars for subjects with fraction times less than (or equal to) and greater than 0.5, respectively. The symbol * indicates *p*-value smaller than 0.05 (Benjamini-Hochberg correction for 9 comparisons). *n* changes for each sub-population and ranges between 38 and 343 (exact values below each boxplot). Panel C. Distribution of normalized volumes for three well-known regions in the sub-populations identified by fraction times of each dFCs as described in Panel B. The symbol * indicates *p*-value smaller than 0.05 (Benjamini-Hochberg correction for 9 comparisons). *n* changes for each sub-population and ranges between 48 and 378 (exact values below each boxplot).

## Discussion

In this study, we introduced a novel framework integrating a Hidden Markov Model (HMM) within a convolutional neural network (CNN) to capture dynamic functional connectivity (dFC) states in subjects across the Alzheimer’s disease (AD) continuum. Our results provide compelling evidence that functional connectivity states do not disappear with disease progression but rather become progressively harder to reach. These findings suggest that the weakening of specific links or the atrophy of brain regions may be a consequence of shifts in the dominant connectivity patterns, rather than the result of isolated or localized damage. While this hypothesis requires further investigation, it opens new directions in understanding AD pathophysiology and the progression of neurodegenerative disorders.

### Dynamic functional connectivity and disease progression

Previous studies have demonstrated that brain networks, particularly the default mode network (DMN) and salience network (SAN), exhibit reduced functional connectivity in AD patients ***Agosta et al. (2012); Brier et al. (2012); Greicius et al. (2004); Dai et al. (2015); Gu et al. (2020***). Traditionally, these changes have been interpreted as localized network disruptions due to structural atrophy, particularly in regions like the hippocampus and posterior cingulate cortex (PCC) ***He et al. (2007); Bullmore and Sporns (2009); Teipel et al. (2018***). However, our findings suggest a different mechanism: the reduced frequency with which certain dynamic states are accessed could underlie the decline in functional connectivity observed in these regions. Rather than being disrupted altogether, these states persist but become harder to reach, implying that network flexibility diminishes over time.

This interpretation is in line with the emerging understanding of AD as a disorder not only of localized damage but also of global network dysfunction. Our results support the idea that the brain’s inability to transition between different connectivity states could drive functional decline, reinforcing the need to study connectivity dynamics as a hallmark of disease progression.

### Compensatory mechanisms and network flexibility

In the early stages of AD, some studies suggest that hyperconnectivity may act as a compensatory mechanism to maintain cognitive performance ***Chiesa et al. (2019); Li et al. (2018); Chen et al. (2020***). Our analysis provides further evidence for this, as subjects with early mild cognitive impairment (MCI) exhibited relatively higher fractions of time spent in cognitively demanding states compared to later stages. These findings align with the two-stage model of AD progression, in which initial compensation is followed by a gradual breakdown of connectivity.

However, the diminishing ability to reach certain states may signify the loss of this compensatory capacity, leading to cognitive decline. The decreased network flexibility observed in AD patients may stem from a progressive inability to transition from states characterized by unimodal sensory network activity to more polymodal, higher-order states. This suggests that targeting treatments to preserve or restore network flexibility could be a promising avenue for early interventions.

### Linking connectivity states to behavior and anatomy

One of the key contributions of this study is the linkage between dynamic connectivity states, cognitive performance, and anatomical atrophy. Our results showed that the fraction of time spent in specific connectivity states correlated with well-established clinical measures such as the Alzheimer’s Disease Assessment Scale (ADAS11) and Mini-Mental State Examination (MMSE), as well as anatomical atrophy in regions like the hippocampus and amygdala. This provides a new perspective on how the brain’s dynamic functional connectivity relates to both behavior and structural degeneration.

While previous studies have explored the relationship between functional connectivity and cognitive decline ***Bergamino et al. (2024); Huang et al. (2022***), few have made direct connections between dynamic states, behavior, and anatomical features. Our findings suggest that the weakening of certain connectivity states is not merely a reflection of structural damage, but may actively contribute to the progression of neurodegenerative changes. These results support the notion that alterations in functional connectivity could precede and potentially drive the structural atrophy observed in key regions affected by AD.

### Implications and future directions

The implications of our findings are twofold. First, they highlight the need to investigate the temporal dynamics of functional connectivity in greater detail, particularly in relation to how these dynamics evolve with disease progression. The persistence of connectivity states, even in advanced AD, suggests that interventions aimed at preserving the brain’s ability to transition between states could help slow cognitive decline. This aligns with recent research advocating for network-based approaches to neurodegeneration, where restoring or maintaining connectivity may be more important than targeting specific regions of atrophy ***Hammond et al. (2020); Strom et al. (2022***).

Second, our work underscores the potential of dynamic connectivity analysis as a diagnostic tool. By identifying patterns of state transitions that correlate with cognitive performance and anatomical degeneration, we can develop more sensitive biomarkers for early-stage AD detection. Additionally, this approach may be applicable to other neurodegenerative conditions, such as Parkinson’s disease or frontotemporal dementia, where similar disruptions in brain network dynamics are thought to play a role.

### Limitations

Despite these promising findings, our approach has several limitations. While we successfully identified connectivity states associated with cognitive decline, the long durations of these states may limit their utility for direct classification purposes. The persistence of states across different stages of AD suggests that classification based on state frequency alone may not capture the full complexity of the disease. Future work should explore more granular, transient sub-states that could provide additional diagnostic value.

Moreover, the link between dynamic connectivity states and structural atrophy, while compelling, requires further validation. Longitudinal studies are needed to determine whether changes in network flexibility precede or follow anatomical degeneration. Additionally, our current model does not account for individual variability in disease progression, which may affect the generalizability of our findings to broader populations.

## Conclusion

This study provides strong evidence for the role of dynamic functional connectivity in Alzheimer’s disease progression. Our results suggest that changes in network flexibility, rather than the disappearance of specific connectivity states, may drive cognitive decline and neurodegeneration. By linking dynamic connectivity states to both cognitive performance and anatomical atrophy, we offer a novel perspective on how brain network dynamics contribute to AD pathology. These findings pave the way for future investigations into network-based interventions that aim to preserve functional flexibility and slow disease progression.

## Material and Methods

### Study cohort

The data used in this study were obtained from the Alzheimer’s Disease Neuroimaging Initiative (ADNI) Phase 3 (ADNI-3) and include T1-weighted and resting-state functional MRI (rs-fMRI) scans. The selected cohort comprises 601 subjects, categorized into five groups: 225 healthy controls (HC), 155 significant memory concern (SMC), 143 early mild cognitive impairment (eMCI), 130 late mild cognitive impairment (lMCI), and 67 individuals with Alzheimer’s disease (AD). Further details on the cohort demographics are available in the Supplementary Information.

### MRI data acquisition and preprocessing

Resting-state fMRI scans were acquired using 3T MRI scanners, with 200 fMRI volumes collected for most participants. T1-weighted structural images were also available for all subjects. Preprocessing of the rs-fMRI data followed standard procedures, including motion correction, normalization, and spatial smoothing. Further details on the acquisition parameters and preprocessing steps are available in the Supplementary Information.

### Deep network model

The deep network model employed in this study is designed to address specific challenges in the analysis of dynamic FC. The structure of this network is presented in the following table ??, along with the dimension of the activations of each layer in space (S), channels (C) and time (T). Further details on each layer can be found in the Supplementary Information.

**Table 5.**
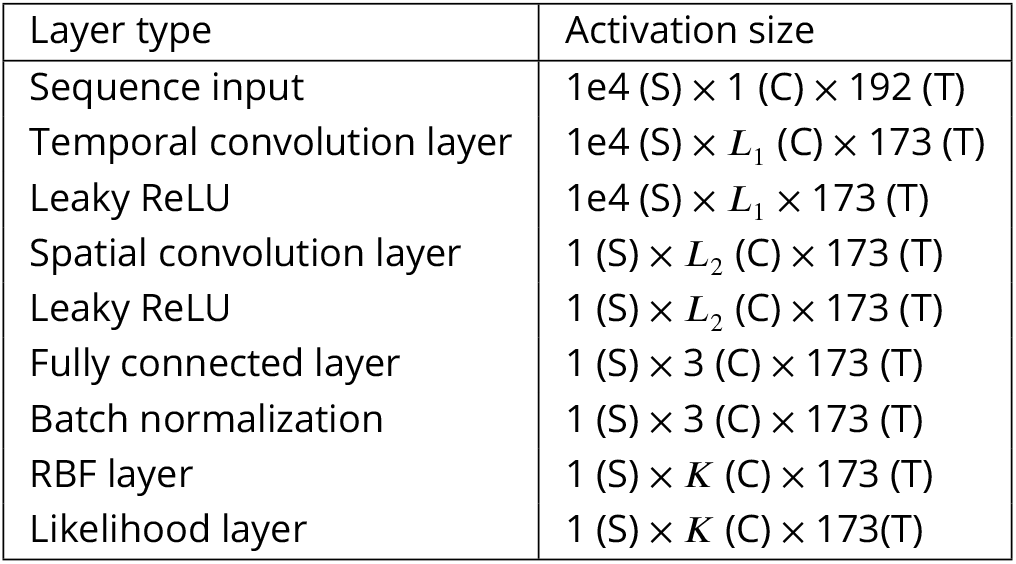
Network model structure and activations

#### Convolutional layers

The convolutional section, inspired by the neural network described in Jie et al. ***Jie et al. (2020***), is designed to overcome the issues associated with the sliding-window approach and identify relevant spatial patterns. This is achieved by designing short-scale time and spatial kernels that provide a compact representation of the input data.

#### Leaky ReLUs

We chose to adopt leaky ReLUs as non-linearities because the radial basis function layer (described below) is sensitive to the distribution of its inputs. Ideally, the inputs to the radial basis function layer should resemble a mixture of several multivariate normal distributions. However, when paired with ReLUs or tanh layers, the inputs are often concentrated around extreme values, causing the radial basis function layer to under-perform. Leaky ReLUs help mitigate this issue by allowing a wider range of values to pass through, resulting in a more suitable input distribution for the radial basis function layer.

#### Radial basis function

A radial basis function (RBF) layer is incorporated to estimate the likelihoods of the outputs from the first section of the network. This layer essentially estimates the probability of the system being in a specific state at a given time, independent of its relation with preceding and following time points.

#### Likelihood layer

The final layer, termed the ’Likelihood layer’, implements the core of the HMM. This layer ensures that the identified states are not just the most likely in isolation, but also when considering the entirety of the time sequence. It computes the likelihood of observing the sequence of inputs, taking into account the temporal dependencies between states.

#### Loss function

The loss function used for network training is a function of two terms: the first is the opposite of the log-likelihood of the most probable sequence, while the second term is obtained by computing the same quantity for a sham sequence in which the time positions of the network inputs have been randomly shuffled. This second term has been introduced to prevent a possible degenerate solution, in which the width parameter of all RBF neurons but one become so small as to be irrelevant and each sequence is decoded as a constant repetition of the state corresponding to the only “large” neuron left. This issue is prevented with the addition of the second loss term, which ensures that the order of inputs is relevant.

### Network training

The optimizer used was ADAM, learning rate was constant and set to 1e-3. Gradient decay was set to 0.75, while squared gradient decay to 0.95. Batch size corresponded to ∼4% of available subjects in the HC-AD experiments, ∼2% when analyzing all available data. Training proceeded for 15 epochs, with no early-stop conditions. *L*_1_, *L*_2_, and *K* were constant for all experiments and set to 16, 32, and 15, respectively. The length of the temporal filters was 20 time volumes, while spatial filters had the same size as the input space (1e4 dimensions).

### Dynamic functional connectivity states analysis

#### Fraction times

The output of the network described above is the clustering of each time volume, for every subject, in one of the possible dynamic functional connectivity states (dFCs). We decided to compare subjects across populations in terms of the fraction times (*f*_*k*_) of each dFCs, given by the percentage of times during which the state is active:

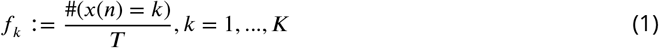

where #(*a*) is the number of times for which condition *a* is verified and *T* is the total length of the time series (in this case, the length of the network output, 173).

#### dFCs sorting and permutation test

The network training procedure is completely unsupervised: as a consequence, the order of clusters is random in each training run. We addressed this problem by sorting the identified dFCs: fraction times for each dFCs were computed and then averaged separately for HC and AD populations. Sorting occurred based on the difference between average fraction time in AD and average fraction time in HC.

An additional issue we encountered is that, due to the stochastic initialization of the network, the attribution of dFCs is not entirely consistent across different training runs. To verify that the observed overlap of dFCs across all training runs is significantly greater than what could be expected by chance alone, we performed a permutation test.

In more detail, we first calculated the total number of time volumes attributed to the same dFCs across all subjects and training runs. Then, we randomly shuffled the dFCs labels and performed the same calculation. This label-shuffling procedure was repeated 10,000 times, generating a distribution of counts under the null hypothesis that the overlap of dFC states is due to chance.

Next, we used a two-sided Wilcoxon signed-rank test to assess whether the median of the distribution of counts from the permutations is significantly different from the observed count of overlapping dFCs. This test allowed us to determine if the observed degree of overlap in dFCs attributions across training runs is greater than what would be expected by chance alone.

#### Fraction times group comparisons

For the dFCs consistently identified across training runs, we assessed whether the observed fraction of time spent in each dFCs significantly differed across the five available populations (HC, SMC, eMCI, lMCI, and AD): we conducted a Kruskal-Wallis test, with the null hypothesis stating that the observed fraction time for each population originates from the same underlying distribution. To account for multiple comparisons and control the family-wise error rate, we applied a Dunn-Sidak correction to the p-values obtained from pairwise comparisons between populations (with a total of 10 pairwise comparisons in this case).

#### Clinico-anatomical comparisons and age correction

Association tests between the fraction times of each dFCs and clinico-anatomical variables were performed only on subjects in the SMC and MCI populations. The rationale for this choice is that both the HC and AD groups were used for the ranking of the identified clusters; thus, their use in this case could produce spurious results.

For each association test, we split the combined SMC-MCI population into two subgroups based on the fraction time of the dFC currently being tested. One subpopulation consisted of subjects who showed the dFC under examination for more than half of the available time points, while the other subpopulation included the remaining subjects. In order to eliminate a possible confound due to age, we used an iterative approach to balance the age distribution between the two supopulations: we randomly selected and removed a subject from the most extreme age quartile from the majority subpopulation until the ages of the two groups were not significantly different, as determined by a Wilcoxon rank test for equal medians with a significance level of 0.2. Finally, we tested each clinical and anatomical variable for different medians among the two subpopulations using a two-sided Wilcoxon rank sum test, followed by a Benjamini-Hochberg correction for multiple tests. dFCs-feature pairs that rejected the null hypothesis of equal medians were considered to have a significant association.

### Hardware and software

All the algorithms described in this paper have been developed on a Asus ROG Strix PC, sporting an AMD Ryzen 9 6900HX and NVIDIA GeForce RTX 3070 Ti Laptop GPU. The OS is Microsoft Windows 11, while the actual code has been developed in MATLAB R2022a.

## Data Availability

Original data is from the ADNI 3 database
Code data is available through github

https://fnih.org/our-programs/alzheimers-disease-neuroimaging-initiative-3-adni-3/

---

**Table S1.** Participants’ demographics

|  | # | Age | Sex (M/F) | APOE4 (0/1/2/U) | ADAS11 | MMSE | CDRSB |
| --- | --- | --- | --- | --- | --- | --- | --- |
| HC | 226 | 71.4 ± 6.1 | 99/127 | 136/50/5/35 | 6.2 ± 4.3 | 28.7 ± 1.8 | 0.37 ± 1.3 |
| SMC | 160 | 71.6 ± 6.0 | 66/94 | 102/51/5/2 | 5.4 ± 3.2 | 29.1 ± 1.2 | 0.11 ± 0.39 |
| eMCI | 144 | 70.9 ± 7.2 | 78/66 | 75/38/13/18 | 8.8 ± 5.7 | 27.7 ± 2.4 | 1.71 ± 2.05 |
| IMCI | 130 | 72.0 ± 8.0 | 72/58 | 63/41/9/17 | 12.1 ± 6.6 | 26.4 ± 4.2 | 2.34 ± 2.59 |
| AD | 67 | 74.8 ± 8.0 | 43/24 | 15/21/10/21 | 20.2 ± 6.4 | 22.7 ± 2.4 | 4.58 ± 2.00 |

**Table S2.** Participants’ demographics

|  | # | Age | Sex (M/F) |
| --- | --- | --- | --- |
| HC | 226 | 71.4 ± 6.1 | 99/127 |
| SMC | 160 | 71.6 ± 6.0 | 66/94 |
| eMCI | 144 | 70.9 ± 7.2 | 78/66 |
| IMCI | 130 | 72.0 ± 8.0 | 72/58 |
| AD | 67 | 74.8 ± 8.0 | 43/24 |

## Supplementary information

### Study Cohort Details

The demographic information of the cohort, including age, sex, and clinical scores, is reported in the tables S1, S2 and S3.

### Clinical and Cognitive Assessments

In this study, several well-established biomarkers and clinical assessments were used to evaluate the subject population. Below is a detailed explanation of each measure:

- **APOE4:** Apolipoprotein E (APOE) is a gene associated with Alzheimer’s disease (AD) risk. The APOE4 allele is considered a major genetic risk factor for late-onset Alzheimer’s disease. Individuals carrying one or two copies of the APOE4 allele are at an increased risk of developing AD.
- **ADAS11:** The Alzheimer’s Disease Assessment Scale-Cognitive Subscale (ADAS-Cog) is a standard cognitive test used to measure the severity of cognitive impairment in Alzheimer’s patients. ADAS11 refers to an 11-item version of this test, focusing on areas such as memory, language, and praxis.
- **MMSE:** The Mini-Mental State Examination (MMSE) is a widely used test for screening cognitive function. It includes questions designed to assess orientation, attention, memory, language, and visual-spatial skills. A lower score on the MMSE indicates greater cognitive impairment.
- **CDRSB:** The Clinical Dementia Rating-Sum of Boxes (CDRSB) is a measure used to assess the severity of dementia symptoms. It provides a global rating of dementia severity by scoring patients across six domains, including memory, orientation, judgment, and personal care.

**Table S3.** *p*-values of age comparisons in different populations, Kruskal-Wallis test for equal distributions, Dunn-Sidak correction for multiple comparisons (*n*=10).

| Group 1 | Group 2 | P-value |
| --- | --- | --- |
| HC | SMC | **0.02** |
| HC | eMCI | 0.87 |
| HC | IMCI | 0.10 |
| HC | AD | **<1e-4** |
| SMC | eMCI | **<1e-2** |
| SMC | IMCI | **<1e-5** |
| SMC | AD | **<1e-9** |
| eMCI | IMCI | 0.64 |
| eMCI | AD | **<1e-2** |
| IMCI | AD | 0.09 |

### MRI Data Acquisition and Preprocessing Details

#### Acquisition parameters

Resting-state fMRI data were acquired on 3T MRI scanners with the following parameters (up-to-date information on the acquisition protocols can be found at https://adni.loni.usc.edu/methods/documents/protocols/).

- Repetition/echo time (TR/TE): 3000/30 ms
- Flip angle (FA): 90°
- Field of view (FOV): 220×220×163 mm
- Voxel size: 3.4-mm isotropic
- Number of volumes: 200 (with small variations, e.g., 197 volumes in a few cases)

T1-weighted structural images were collected with the following parameters:

- TR = 2300 ms, TE = minimum
- Inversion time (TI) = 900 ms
- FOV = 208×240×256 mm
- Voxel size: 1-mm isotropic

#### Preprocessing Steps

Resting-state fMRI data were preprocessed using the FMRIB Software Library (FSL version 6.0) with the following steps:

- **Initial preprocessing:**
  - Removal of the first 5 volumes
  - Motion correction using MCFLIRT
  - 4D mean intensity normalization
  - Spatial smoothing with a 6-mm full-width half maximum (FWHM) kernel
  - Interleaved slice-timing correction
- **Regressing out confounds:**
  - The six motion parameters (plus their derivatives), white matter (WM), and cerebrospinal fluid (CSF) signals were regressed out along with a linear trend component.
  - WM and CSF signals were extracted from segmented T1-weighted scans, registered to the fMRI native space, and eroded and binarized with a threshold of 0.8.
  - **Band-pass filtering:**
  - A band-pass filter was applied with a frequency range of 0.01-0.08 Hz.
- **Scrubbing and zero-padding:**
  - High-motion frames (defined as exceeding 0.5 mm framewise displacement) were scrubbed.
  - Zero-padding of the volumes, along with one preceding and two subsequent volumes, was performed to ensure data consistency across subjects.
- **Normalization:**
  - The preprocessed rs-fMRI volumes were spatially normalized to the 2-mm Montreal Neurological Institute (MNI) space using both linear and non-linear registration techniques (FLIRT and FNIRT).

The Schaefer functional atlas ***Schaefer et al. (2018***) with 100 parcels and 7 RSNs was used to extract the mean time course for each region of interest (ROI), which were normalized using *Z*-score before further processing.

The T1-weighted volumes were minimally preprocessed for bias-field correction (*fsl_anat* tool ***Jenk- inson et al. (2012***)), and a complete brain parcellation/segmentation was performed using FreeSurfer version 7.0 ***Fischl (2012***). In this study, we focused our analyses on three regions of interest, that are Hippocampus and Amygdala, which are relevant to AD, and cerebellum cortex, which is a control region that is not typically affected by the disease ***Braak and Braak (1991); Thompson et al. (2003); Frisoni et al. (2002***). The volumes of these regions were then normalized by the estimated total intracranial volume of the respective subject, and averaged over hemispheres.

### Network structure

#### Convolutional layers

The input of the network is the vectorized, pairwise product of the timeseries for all ROIs, while the shape of the time window used to compute the correlations is learned by the network itself (i.e., it is defined by the kernels of the time convolution layer of the network). More in detail:

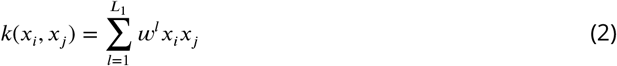

where *x*_*i*_ and *x*_*j*_ are the normalized time series for ROIs *i* and *j*, while 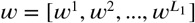 is one of the learnable weight vectors. The proposed approach reduces to the computation of Pearson’s coefficient of correlation if all elements in *w* equal 1. Differently from ***Jie et al. (2020***), here only one spatial convolutional layer is used. As the dimension of the kernels of this layer equals that of its inputs and no padding is performed, the outputs of this layer project the dFC matrices computed with the windowing technique described above to a space of dimension 1 ×*L*_2_. Here, *L*_1_ and *L*_2_ are the number of kernels used in the temporal and spatial convolutional layer, respectively.

#### Non-linearity layers

Each convolutional layer described above is followed by a leaky ReLU layer with scale parameter *α* = 0.5. The element-wise transformation applied by this layer is thus described by:

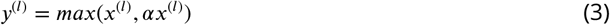

#### Fully connected and batch normalization layer

The fully connected layer is added to perform a final projection of each input to a 1 × 3 vector. The set of experiments we performed showed that increasing this dimension resulted in additional computation time with no significant differences in terms of final output results (data not shown).

A batch normalization layer has been added to prevent degenerate solutions: as the network is trying to maximize the likelihood of the input sequences (see), an obvious (and non-useful) solution would be to collapse all data to the same value, e.g., by setting all the weights of the fully connected layer to zero. The presence of a batch normalization layer prevents that from happening, as it decouples network loss from simple scale manipulations that would eventually lead to a degenerate solution.

#### Radial basis function

Each radial basis function neuron, as implemented here, learns the parameters of a multivariate normal distribution with covariance matrix Σ = *σ*^2^*I*.

*x*^*s*^(*t*) is the 1 × 3 output of the preceding fully connected layer for subject *s* at time *t* and the output 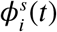 is the likelihood of *x*^*s*^(*t*) for hidden state *i*:

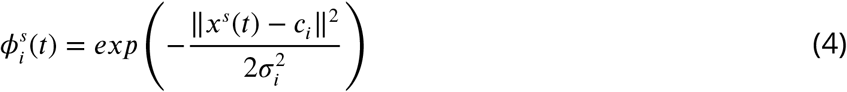

where ‖ · ‖ is the Euclidean norm, *c*_*i*_ and *σ*_*i*_ are the learned parameters of neuron *i* (center and width, respectively).

#### Likelihood layer

The last layer receives as inputs the sequence in time of likelihoods for each state and provides as output the likelihood of the most probable time sequences. This layer implements part of the Viterbi algorithm: all states have the same probability for *t* = 1, while each element of the transition matrix *A*^*s*^ for subject *s* is estimated as:

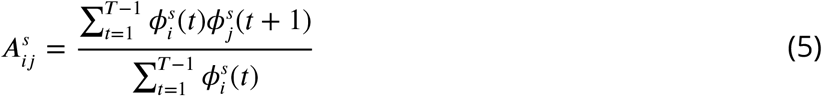

The transition matrix *A* is estimated at each iteration as the average of *A*^*s*^ across all subjects. 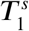 at the last time point contains the likelihood of the observations for the most likely sequence of states for subject 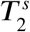 contains the information needed to reconstruct the most likely sequence of visited hidden states. However, the sequence itself is not required to compute the loss function (see Section) and it is therefore computed separately from network operations.

#### Loss function

The loss function used for network training is a function of two terms: the opposite of the log-likelihood of the most probable sequence (i.e., *L*_*abs*_ = −*log*(*max*_*i*_(*T*_1_[*i, T* ])) and the same quantity for a sham sequence *L*_*sham*_ in which the time positions of the network inputs have been randomly shuffled.

The definition of the complete loss term used in training is, therefore:

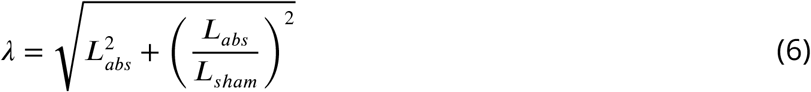

